# Brain Total Creatine Differs Between Primary Progressive Aphasia (PPA) Subtypes and Correlates with Disease Severity

**DOI:** 10.1101/2022.05.20.22275384

**Authors:** Kathleen E. Hupfeld, Helge J. Zöllner, Georg Oeltzschner, Hayden W. Hyatt, Olivia Herrmann, Jessica Gallegos, Steve C. N. Hui, Ashley D. Harris, Richard A. E. Edden, Kyrana Tsapkini

**Author notes:** **Corresponding Author:** Kyrana Tsapkini, PhD, Departments of Neurology and Cognitive Science, Johns Hopkins University, Phipps 488, 600 N Wolfe Street, Baltimore, MD 21287, <u>Email</u>.

## Abstract

Primary progressive aphasia (PPA) is comprised of three subtypes: logopenic (lvPPA), non-fluent (nfvPPA), and semantic (svPPA). We used magnetic resonance spectroscopy (MRS) to measure tissue-corrected metabolite levels in the left inferior frontal gyrus (IFG) and right sensorimotor cortex (SMC) from 61 PPA patients. We aimed to: 1) characterize subtype differences in metabolites; and 2) test for metabolite associations with symptom severity. tCr differed by subtype across the left IFG and right SMC. tCr levels were lowest in lvPPA and highest in svPPA. tCr levels predicted lvPPA versus svPPA diagnosis. Higher IFG tCr and lower Glx correlated with greater disease severity. As tCr is involved in brain energy metabolism, svPPA pathology might involve changes in specific cellular energy processes. Perturbations to cellular energy homeostasis in language areas may contribute to symptoms. Reduced cortical excitatory capacity (i.e., lower Glx) in language regions may also contribute to symptoms. Thus, tCr may be useful for differentiating between PPA subtypes, and both tCr and Glx might have utility in understanding PPA mechanisms and tracking progression.

**Highlights:**

- No prior work has examined differences in brain metabolite levels for PPA subtypes.
- Total creatine (tCr) levels were lowest in lvPPA and highest in svPPA.
- tCr levels differentiated lvPPA from svPPA diagnosis.
- Higher tCr and lower Glx in the left IFG correlated with greater disease severity.
- Changes in cellular energy and excitatory processes may relate to PPA pathology.

## 1. Introduction

Primary progressive aphasia (PPA) is a devastating neurodegenerative condition characterized by prominent declines in language function (Gorno-Tempini et al., 2011; Mesulam, 1982). Symptoms usually begin in middle age (around 40-60 years), worsen over time, and may later extend to other cognitive domains (Gorno-Tempini et al., 2011; Grossman, 2010; Mesulam, 1982). PPA thus considerably impacts quality of life, in addition to shortening life expectancy; average life expectancy after symptom onset is only about 7-12 years (Tastevin et al., 2021). Critically, aside from some symptom improvement with behavioral therapy (Pagnoni et al., 2021) or neuromodulation (Cotelli et al., 2020; Tsapkini et al., 2018), there remain no disease-modifying treatments for PPA. Therefore, it is becoming increasingly imperative to improve scientific understanding of PPA pathology to identify biomarkers of disease progression as well as potential therapeutic targets.

Consistent with their language symptoms, those with PPA typically display cortical atrophy in the left hemisphere, particularly in brain regions related to language function (Rogalski et al., 2014). However, PPA is a heterogeneous disorder associated with both varied symptoms and varied pathology. As such, expert consensus has divided PPA into three main subtypes (Gorno-Tempini et al., 2011) (along with mixed or unclassified cases): 1) logopenic variant PPA (lvPPA) is characterized by deficits in word retrieval (i.e., producing the intended word) and difficulty repeating words and sentences; 2) non-fluent variant PPA (nfvPPA) is characterized by deficits in grammar and speech production and/or slow, effortful, and distorted (apraxic) speech, but preserved word comprehension and naming; and 3) semantic variant PPA (svPPA) is characterized by severe deficits in single-word comprehension and naming, but preserved grammar and fluency (i.e., these individuals tend to produce “empty” speech).

These three subtypes have been linked to different cortical atrophy patterns and different pathology. Atrophy is most prominent in the left posterior temporal and inferior parietal lobes in lvPPA, left posterior frontal and fronto-insular cortex in nfvPPA, and anterior temporal lobes (left dominant but usually bilateral) in svPPA (Gorno-Tempini et al., 2011). LvPPA is most often associated with Alzheimer’s disease (AD) pathology (e.g., PET-PIB positivity, decreased Aβ42, and elevated tau in the cerebrospinal fluid) (Mesulam et al., 2008; Rabinovici et al., 2008), nfvPPA is most often associated with tau-positive pathology (Josephs et al., 2006; Knibb et al., 2006; Mesulam et al., 2008), and svPPA is most often associated with ubiquitin/TDP43-positive frontotemporal lobar degeneration (Davies et al., 2005; Grossman, 2010; Hodges et al., 2004; Mesulam et al., 2008; Nestor et al., 2007). However, there is not a direct correspondence between atrophy or pathology and PPA subtype; thus, further work is warranted to better understand how disease mechanisms might differ between the PPA subtypes.

Magnetic resonance spectroscopy (MRS) permits measurement of brain chemicals *in vivo* in humans using an MRI scanner and thus can provide insight into potential disease mechanisms and drug targets, distinct from conclusions that can be drawn from the atrophy and pathology measurements discussed above. Standard MRS acquisitions, including short-echo time (TE) Point RESolved Spectroscopy (PRESS), measure metabolites including total *N*-acetyl aspartate (tNAA), total choline (tCho), total creatine (tCr), and glutamate+glutamine (Glx) (Bottomley, 1982). tNAA, a composite measure of NAA and NAAG (N-acetylaspartylglutamate), is a neuronal marker, suggested as an indicator of neuronal density and integrity (Rae, 2014). tCho, the composite of phospholipid precursors phosphocholine (PCh) and glycerophosphocholine (GPC), can reflect changes in membrane turnover or cell density (Cleeland et al., 2019). Creatine and phosphocreatine (reported in combination as tCr) are involved in brain energy metabolism and homeostasis (Rae, 2014). Glx is a combination of two signals: glutamate (i.e., the principal excitatory neurotransmitter in the central nervous system) and glutamine (i.e., a precursor for glutamate). *J*-difference editing of the spectrum using MEscher–GArwood Point RESolved Spectroscopy (MEGA-PRESS) (Choi et al., 2021; Harris et al., 2017; Mescher et al., 1998) allows us to quantify additional metabolites with coupled spin systems and overlapped signals. These include the primary inhibitory neurotransmitter within the central nervous system, gamma-aminobutyric acid (GABA).

A majority of studies suggest that brain tNAA, Glx (or glutamate), and GABA levels decrease with normal aging (Chiu et al., 2014; Cleeland et al., 2019; Gao et al., 2013; Haga et al., 2009), and are decreased in mild cognitive impairment (MCI) (Oeltzschner et al., 2019; Zeydan et al., 2017) and AD (Bai et al., 2015; Rupsingh et al., 2011; Wang et al., 2015; Zhang et al., 2014) compared with normal aging. Both tCho and tCr either increase or show no change with normal aging (Chiu et al., 2014; Cleeland et al., 2019; Haga et al., 2009), and are either elevated (Huang et al., 2001; Marjańska et al., 2019) or not different (Marjańska et al., 2019; Wang et al., 2015) in MCI and AD compared with normal aging. Functionally, lower MRS-measured tNAA, Glx (or glutamate), and GABA, and higher tCho, have each been associated with poorer cognitive performance in normal aging (Erickson et al., 2012; Gomar et al., 2014; Kantarci et al., 2011; Porges et al., 2017a; Ross et al., 2005), MCI (Griffith et al., 2007; Lim et al., 2012; Oeltzschner et al., 2019; Riese et al., 2015), AD (Lim et al., 2012), and frontotemporal lobar degeneration patients (Murley et al., 2022). Taken together, it is plausible that these metabolites might differ between the three PPA subtypes and associate with disease severity.

We previously identified GABA changes in the left inferior frontal gyrus (IFG) following transcranial direct current stimulation (tDCS) combined with language therapy in PPA patients (Harris et al., 2019). However, the sample size included in this prior work (*n* = 22) was not sufficient to stratify metabolite levels by the three PPA subtypes. Therefore, the objectives of the present study included: 1) to characterize differences in brain metabolite levels between the three PPA subtypes and determine if brain metabolite levels can discriminate between PPA subtypes; and 2) to examine relationships between brain metabolite levels and PPA symptom severity, as indicated by the FrontoTemporal Dementia Clinical Dementia Rating Scale (FTD-CDR, a measure of overall disease severity, including language function, memory, attention, and independence, with scores ranging from 0 (no impairment) to 24 (severe impairment) (Knopman et al., 2008)). We measured MRS metabolites in the left IFG and right sensorimotor cortex (SMC). We selected the left IFG due to its involvement in several language functions such as lexical selection and production, as well as other cognitive functions (Liakakis et al., 2011), evident atrophy in PPA (particularly lvPPA and nfvPPA but also svPPA) (Preiß et al., 2019; Rogalski et al., 2011), and response to tDCS therapy across PPA subtypes (Harris et al., 2019; Tsapkini et al., 2018, 2014). We selected the right SMC (which is involved in sensory processing and motor execution) as a control region. That is, we predicted that brain metabolite levels would differ between PPA subtypes and associate with FTD-CDR scores for the left IFG but not the right SMC.

## 2. Methods

### 2.1 Participants

Prior to study enrollment, all patients were evaluated using neurological examination, cognitive and language testing, and neuroimaging. All patients enrolled in the study were diagnosed with PPA using current consensus criteria (Gorno-Tempini et al., 2011); diagnoses were confirmed by expert clinicians. Additional inclusion criteria included: at least a 12^th^ grade education, right-handedness, and English as a first language. Participants were excluded if: they were over 90 years of age, were not pre-morbidly proficient spellers, had progressed to advanced stages of PPA or other dementia, had any diagnosed comorbid neurologic conditions (e.g., stroke, psychiatric, or developmental disorder), or if they had any contraindications for MRI scanning. The Johns Hopkins University Institutional Review Board (#NA_00071337) approved all study procedures, and the trial was registered on ClinicalTrials.gov (NCT02606422). All participants scored >0 on the FTD-CDR scale, indicating at least some level of impairment to daily functioning. All participants also completed the Boston Naming Test (BNT) (Kaplan et al., 2001), Hopkins Assessment on Naming Actions (HANA) (Breining et al., 2015), digit span (forward and backward), Letter Verbal Fluency Test (FAS), Semantic Verbal Fluency Test (fruits, animals, and vegetables), and the Trail Making Test A and B (Tombaugh, 1999). Of note, the present work included only PPA patients and not age-matched controls; we expand on the limitations of this study design in the Discussion.

### 2.2 MRS Acquisition

All MRS data were collected using the same 3.0 Tesla Philips Achieva MRI scanner using a 32-channel head coil. For voxel positioning and tissue segmentation, we first collected a whole-brain *T*_1_-weighted structural MRI scan, using the following parameters: magnetization-prepared rapid gradient-echo (MPRAGE) sequence, TR/TE = 8 ms/3.75 ms, flip angle = 8°, slice thickness = 1.0 mm, 150 slices, voxel size = 1 mm^3^ isotropic voxels. Next, immediately after the *T*_1_, we acquired metabolite spectra from two 30 × 30 × 30 mm^3^ voxels (Figure 1) using both short-TE PRESS and MEGA-PRESS sequences. We positioned the left IFG voxel using the anatomical landmarks of the lateral ventricles and insula. We positioned the right SMC voxel by centering the voxel on the hand knob in the precentral gyrus. Both voxels were carefully shifted and rotated to avoid the edge of the brain and ventricles as much as possible to avoid lipid contamination and partial-volume effects.

**Figure 1.**
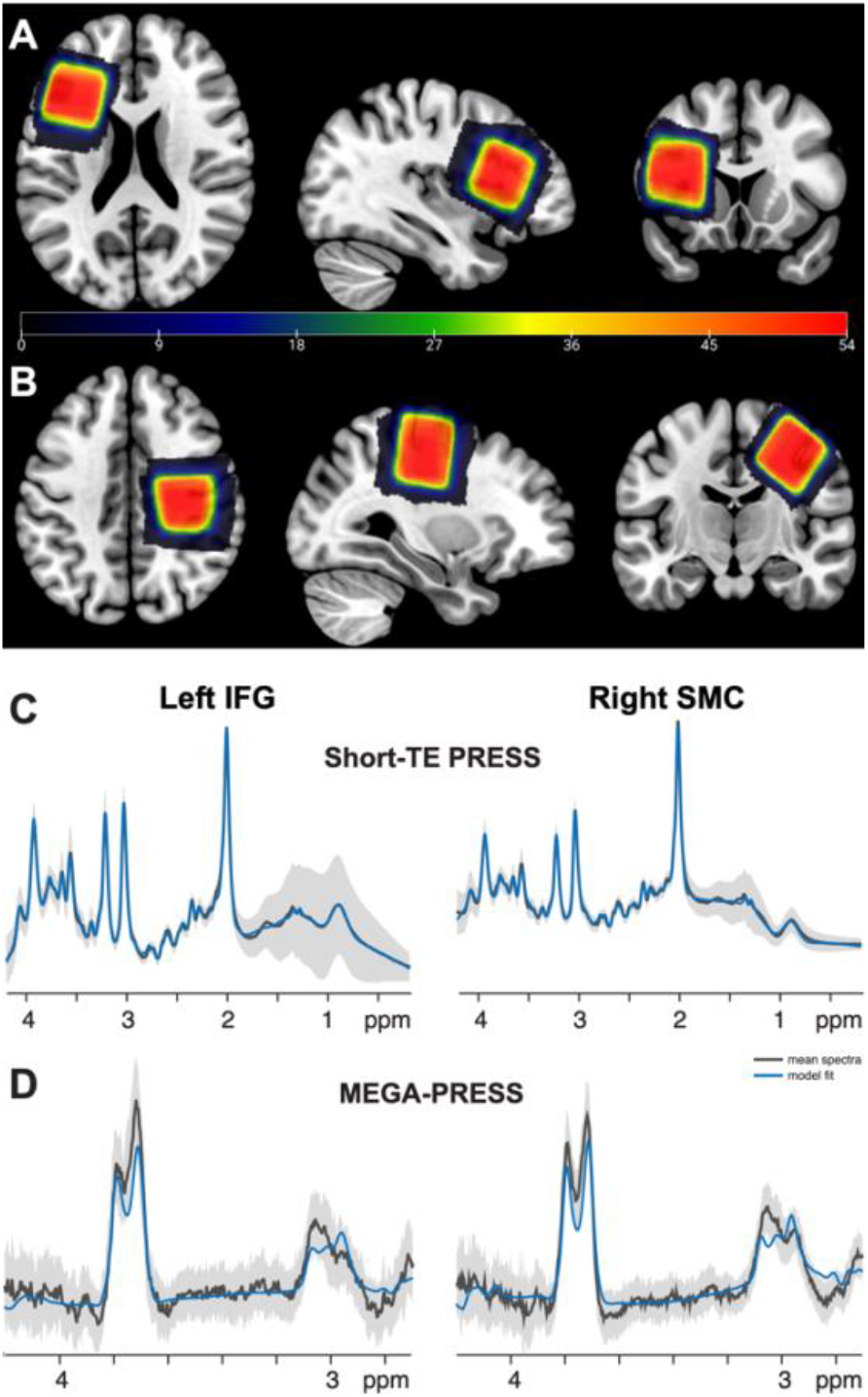
Voxel Placement and Spectra. **(A-B)** *In vivo* MR spectra were acquired from the left inferior frontal gyrus (IFG) and right sensorimotor cortex (SMC); each participant’s native space binary voxel mask is normalized to standard (MNI) space and overlaid onto the spm152 template. Warmer colors indicate areas of greater overlap between participants (color bar = number of subjects overlapped). **(C-D)** Average short-TE PRESS and GABA-edited MEGA-PRESS spectra (black), 95% confidence interval (gray shading) and model fits (blue) for all participants for both voxels. The PRESS spectra are shown from 0.2-4.2 ppm. The MEGA-PRESS spectra are shown from 2.7-4.2 ppm.

To measure tNAA, tCho, tCr, and Glx, we used a standard short-TE PRESS acquisition with the following parameters: TR/TE = 2000 ms/32 ms; 48 averages sampled at 2000 Hz with 2048 points; and water suppression using the VAPOR method (Tkáč et al., 1999). To resolve GABA, we used MEGA-PRESS (Harris et al., 2017; Mescher et al., 1998) with the following parameters: TR/TE = 2000 ms/68 ms; 14-ms editing pulses at 1.9 ppm and 7.46 ppm, alternating every 2 averages; 320 averages in total; and VAPOR water suppression (Tkáč et al., 1999). MEGA-PRESS editing schemes for the detection of GABA co-edits homocarnosine and macromolecular signals (Harris et al., 2015a); therefore, as is standard practice, we refer to the edited 3 ppm GABA signal as GABA+ in the remainder of the manuscript. For both the short-TE PRESS and MEGA-PRESS acquisitions, we also collected 8 water reference spectra without water suppression or pre-inversion, for later use in eddy current correction and metabolite quantification.

### 2.3 MRS Data Processing

MRS data were analyzed using the open-source analysis toolbox Osprey (v2.0.0; https://github.com/schorschinho/osprey/) (Oeltzschner et al., 2020) within MATLAB R2021b. The analysis procedures followed consensus-recommended processing guidelines for PRESS and MEGA-PRESS data (Choi et al., 2021; Wilson et al., 2019) and match those applied in our previous work (Zöllner et al., 2022, 2021). Briefly, for the short-TE PRESS data, analysis steps included: loading the vendor-native raw data (which had already been coil-combined, eddy-current-corrected, and averaged on the scanner at the time of data collection), removing the residual water signal using a Hankel singular value decomposition (HSVD) filter (Barkhuijsen et al., 1987), and modeling the metabolite peaks as described in detail in (Oeltzschner et al., 2020; Zöllner et al., 2021) using a custom basis set generated by our new MRSCloud tool (https://braingps.anatomyworks.org/mrs-cloud; (Hui et al., 2022)). The basis set was generated using PRESS localization at TE = 32 ms with the Philips-specific sequence timing and real RF pulse waveforms, and consisted of simulations of 32 metabolites including NAA and NAAG (=tNAA), Cr and PCr (=tCr), GPC and PCh (=tCho), Glu and Gln (=Glx), and GABA (which are of interest in this study), as well as 23 other low-concentration metabolites. We then created a binary mask of the two MRS voxels in subject space, co-registered these masks to each participant’s *T*_1_-weighted structural scan, and segmented the structural scans using SPM12 (Ashburner et al., 2014), and quantified metabolite levels with respect to the unsuppressed water scan. These tissue volume fractions in the MRS voxels were used for full tissue- and relaxation-corrected metabolite quantification (see below).

For the GABA-edited MEGA-PRESS data additional, standardized analysis steps in Osprey included: eddy-current correction based on the water reference (Klose, 1990), robust spectral registration (Mikkelsen et al., 2020) to separately align the individual transients within each sub-spectrum, and final alignment of the averaged sub-spectra by minimizing the choline peak in the difference spectrum before generating the final GABA-edited difference spectrum. The co-edited macromolecules (MMs) at 3 ppm were modelled with the “1to1GABA” model, which uses one composite GABA + MM basis function (i.e., the sum of the GABA and MM_3co_ basis functions, with a fixed 1:1 amplitude ratio); this model assumes 50% of the 3-ppm signal in the GABA-edited difference spectrum can be attributed to co-edited MMs (Deelchand et al., 2021; Zöllner et al., 2022). As is standard for 3 T GABA-edited MEGA-PRESS data, we considered the composite edited 3-ppm signal, i.e., “GABA+” (GABA+MM) values in our statistical analyses.

Finally, we corrected all metabolite estimates in order to assess metabolite levels only in the tissue present in the voxel, which is particularly relevant for aging and neurodegenerative disease populations who may have substantial cortical atrophy (Maes et al., 2018; Porges et al., 2017b). We applied tissue and relaxation correction to the short-TE PRESS values; this adjusts metabolite estimates to account for atrophy and heterogeneous tissue composition within the voxels, using literature values (Wansapura et al., 1999) to account for differences in water visibility and relaxation times between tissue types (Gasparovic et al., 2006; Harris et al., 2015b). Similarly, to account for the effects of atrophy and tissue-specific water visibility and relaxation, (and assuming that GABA+ levels in gray matter are twice that in white matter), we applied the “alpha” correction to all GABA+ values (Harris et al., 2015b). For further details on MRS data acquisition, processing, and quality, see Appendices A-B, in which we list all consensus-recommended parameters (Lin et al., 2021). For further information on metabolite quantification and the applied tissue correction, see Appendix C.

### 2.4 Statistical Analyses

We conducted all statistical analyses using R 4.0.0 (R Core Team, 2021) within RStudio (RStudio Team, 2021). First, we used one-way ANOVAs (or Kruskal-Wallis non-parametric tests if the ANOVA assumptions of normality and heteroskedasticity were not met) to test for group differences in demographic variables, symptom severity, cognitive function, and MRS data quality between the three subtypes. Next, we used linear mixed models (*lme*, (Pinheiro and Bates, 2000)) to examine whether metabolite levels differed by PPA subtype or brain region. In the final model, we entered the metabolite level as the outcome variable, PPA subtype and voxel (i.e., IFG or SMC) as predictors, and included a random intercept for each subject (similar to our previous work, (Hupfeld et al., 2021)). Including the PPA subtype*voxel interaction term or a random slope for each subject did not improve model fit (likelihood ratio test *p* > 0.05), so these terms were omitted from the final statistical models. Including age or years since diagnosis as additional predictors in follow-up models did not influence the statistical significance of any results. Each final model satisfied the linear mixed model assumptions of homogeneity of variances and normality of residuals. We used this same linear mixed model approach to test for PPA subtype and brain region differences in bulk tissue composition (i.e., voxel gray matter, white matter, and cerebrospinal fluid fractions). We conducted a follow-up analysis using two binary logistic regressions to determine whether tCr levels significantly predicted diagnosis (i.e., lvPPA vs. svPPA and lvPPA vs. nfvPPA). Including age or years since diagnosis in these logistic regression models resulted in poorer model fit (higher AIC and BIC) and thus these two predictors were excluded from the final model.

Lastly, we applied a linear model to test for relationships between brain metabolite levels and PPA symptom severity (i.e., FTD-CDR scores). Given the subgroup sizes and the missing MRS and FTD-CDR data (see Table B2), we conducted this analysis across the whole cohort instead of stratified by PPA subtype. In addition, given the amount of missing GABA+ data compared with the other metabolites (Table B2), we did not include GABA+ as a predictor in this metabolite-symptom severity model (though we did test for GABA+ differences between PPA subtypes, as described above). Thus, in the full model, we entered as predictors: the metabolite levels from both voxels (4 metabolites/voxel), age, sex, and years since diagnosis. We then selected a final model using *bestglm* with cross-validation, delete-d method, and 1,000 replications to produce a final model that retained only the best predictors of FTD-CDR score (McLeod and Xu, 2010; Zhang, 2016). The FTD-CDR scores were not normally distributed (Shapiro test *p* < 0.05), so we reran the same model selection procedure after square root-transforming the FTD-CDR variable, which improved the normality of the data (Shapiro test *p* > 0.05).

## 3. Results

### 3.1 Sample Characteristics

The total cohort consisted of 61 PPA patients: 22 lvPPA, 27 nfvPPA, and 12 svPPA. There were no differences in sex, age, time since diagnosis, or symptom severity based on PPA subtype (Table 1). As anticipated (Beales et al., 2019; Gorno-Tempini et al., 2004; Wicklund et al., 2014), svPPA participants performed worst on the tests of naming (i.e., BNT and HANA) and semantic verbal fluency (Fruits, Animals Vegetables), and lvPPA participants performed worst on the Digit Span – Forward and Backward tasks, which assess verbal working memory (Table 1). There were no subtype differences for the remaining cognitive tasks (i.e., FAS or Trail Making – A and B tests; Table 1). IFG short-TE PRESS data was not collected for 2 participants, IFG MEGA-PRESS data was not collected for 1 participant, and FTD-CDR scores were missing for 7 participants. Table B2 details additional data omissions from statistical analyses; MRS data were excluded if NAA linewidth was greater than 15 Hz (Cao et al., 2018; Deelchand et al., 2021; Mosser and Edwards, 2008; Nwaroh et al., 2020; Sasaki et al., 2021) or if incorrect pulse sequence parameters were used during acquisition. Group mean spectra (after these exclusions) are presented in Figure 1, and spectra for a single exemplar participant are shown in Figure B1. Data quality metrics (i.e., linewidth, SNR, fit error, and drift) are presented for the whole cohort (Appendix A) and by PPA subtype (Appendix B). There were no differences in MRS data quality based on PPA subtype (Table B1).

**Table 1.** Sample Characteristics

Note.* For each metric, we report the mean (standard deviation) for each PPA subtype.
| | lvPPA | nfvPPA | svPPA | F or $\chi^2$ | p | $\eta^2$ |
| --- | --- | --- | --- | --- | --- | --- |
| n | 22 | 27 | 12 | -- | -- | -- |
| Sex | 13 F; 9 M | 11 F; 16 M | 7 F; 5 M | 1.97 | 0.373 | -- |
| Age (years) | 65.55 (8.12) | 68.59 (6.62) | 68.67 (5.25) | 1.36 | 0.265 | 0.04 |
| Years of education<sup>a,b</sup> | 16.48 (1.91) | 16.83 (2.44) | 16.35 (2.33) | 0.23 | 0.799 | 0.01 |
| Years since diagnosis<sup>a,c</sup> | 3.93 (2.83) | 3.81 (2.58) | 5.05 (2.91) | 0.83 | 0.442 | 0.03 |
| FTD-CDR sum score<sup>a,d</sup> | 6.55 (5.17) | 5.26 (4.17) | 7.61 (5.59) | 0.87 | 0.424 | 0.03 |
| Boston Naming Test (BNT)<sup>a,e</sup> | 19.19 (9.03) | 22.00 (8.09) | 4.00 (3.95) | 20.68 | < 0.001*** | 0.43 |
| Hopkins Assessment on Naming Actions (HANA)<sup>a,f</sup> | 20.90 (9.72) | 24.28 (9.62) | 9.10 (6.74) | 9.72 | < 0.001*** | 0.27 |
| Letter Verbal Fluency (FAS)<sup>a,h</sup> | 18.77 (12.38) | 16.58 (10.22) | 21.90 (15.44) | 0.73 | 0.487 | 0.03 |
| Semantic Verbal Fluency (Fruits, Animals, Vegetables)<sup>h</sup> | 20.73 (13.48) | 22.00 (12.75) | 10.10 (7.40) | 3.55 | 0.036* | 0.11 |
| Digit Span – Forward<sup>g</sup> | 3.82 (1.82) | 4.58 (1.64) | 6.35 (1.76) | 7.36 | 0.001** | 0.21 |
| Digit Span – Backward<sup>g</sup> | 2.36 (1.15) | 2.96 (1.24) | 4.80 (1.70) | 12.29 | < 0.001*** | 0.31 |
| Trail Making Test - A<sup>a,i</sup> | 98.18 (97.26) | 53.96 (21.14) | 47.89 (17.90) | 3.14 | 0.053 | 0.12 |
| Trail Making Test - B<sup>a,i</sup> | 233.12 (85.03) | 196.11 (89.90) | 147.89 (96.74) | 2.71 | 0.078 | 0.11 |

<sup>a</sup> In cases where the variable did not meet the one-way ANOVA assumptions of homogeneity of variances between groups, independence of residuals, and/or normality of residuals (indicated by the subscript 'a'), we reran each analysis using the non-parametric Kruskal-Wallis test. In all cases, the non-parametric test did not change the statistical significance of the result.
<sup>b</sup> Years of education was unavailable for 1 lvPPA, 3 nfvPPA, and 2 svPPA patients.
<sup>c</sup> Years since diagnosis was unavailable for 1 lvPPA, 3 nfvPPA, and 1 svPPA patients.
<sup>d</sup> FTD-CDR sum score was unavailable for 4 nfvPPA patients and 3 svPPA patients.
<sup>e</sup> BNT was scored out of 30 possible points. BNT score was unavailable for 1 lvPPA, 2 nfvPPA, and 1 svPPA patient.
<sup>f</sup> HANA was scored out of 35 possible points. HANA score was unavailable for 1 lvPPA, 2 nfvPPA, and 2 svPPA patients.
<sup>g</sup> Digit Span – Forward and Backward were scored out of 9 possible points each. Digit span scores were unavailable for 1 nfvPPA patient and 2 svPPA patients.
<sup>h</sup> FAS and Fruits, Animals, Vegetables were scored as the number of words the participant could name starting with the letters F, A, and S or in each fruit/animal/vegetable category (1 minute for each trial). These scores were unavailable for 1 nfvPPA patient and 2 svPPA patients.
<sup>i</sup> Trail Making Test – A and B scores are in seconds required to complete the task. Trail Making Test – B was terminated after 300 seconds. Trail Making Test scores were unavailable for 5 lvPPA, 6 nfvPPA, and 3 svPPA patients.

### 3.2 Differences in Brain Metabolite Levels by PPA Subtype

tCr levels differed by subtype across both voxels; svPPA patients had higher mean tCr compared with lvPPA patients (*p* = 0.019; Figure 2; Table 2). nfvPPA patients had higher mean tCr levels compared with lvPPA patients, though this difference did not reach statistical significance within the linear mixed effects model (*p* = 0.071; Figure 2; Table 2). Including age in this model did not change the statistical significance of any results (svPPA vs. lvPPA: *p* = 0.045; nfvPPA vs. lvPPA: *p* = 0.175). However, age was also a significant predictor of tCr levels; older age was associated with greater tCr levels (*p* = 0.011). There were no other differences in brain metabolite levels (all *p* > 0.05; Figure 2; Table 2) or voxel tissue fractions (all *p* > 0.05; Table B4) based on PPA subtype. Moreover, there were no associations between years since diagnosis and any metabolite levels (all *p* > 0.05).

**Figure 2.**
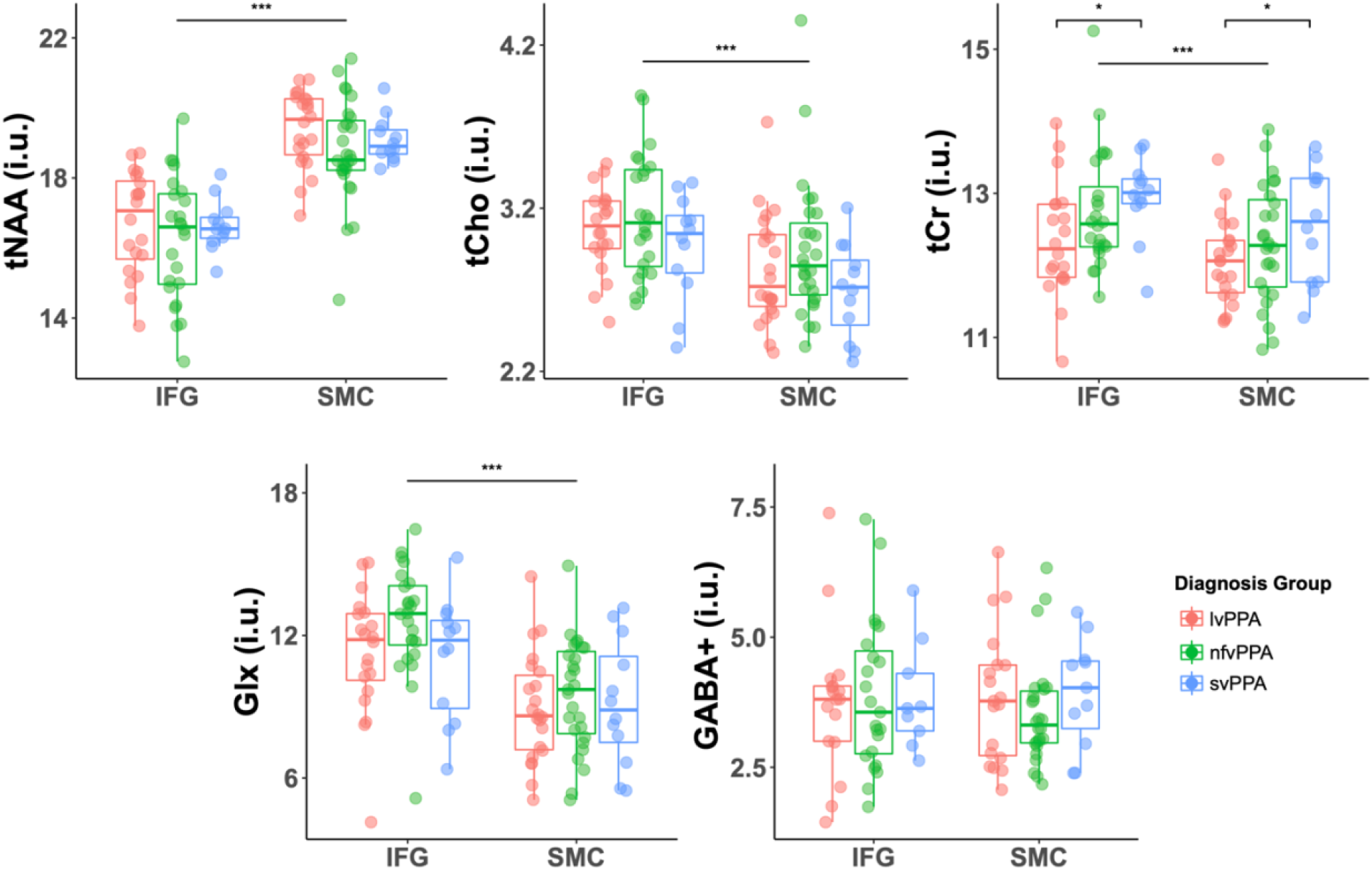
Differences in Metabolites by PPA Subtype and Brain Region. Metabolite levels for the lvPPA (pink), nfvPPA (green), and svPPA (blue) subtypes are shown for the IFG (left) and SMC (right) voxels. * indicates *p* < 0.05 for the comparison of svPPA tCr levels with lvPPA tCr levels. *** indicates *p* < 0.001 for the comparison of metabolite values within the SMC versus the IFG.

**Table 2.** Differences in Metabolites by PPA Subtype and Brain Region

Note. Here we report the results of a linear mixed effects model testing for PPA subtype and voxel differences in each metabolite.
| Predictors | Estimates (SE) | CI | t | p |
| --- | --- | --- | --- | --- |
| **tNAA (i.u.)** |  |  |  |  |
| Diagnosis (nfvPPA) | -0.53 (0.33) | -1.19-0.12 | -1.60 | 0.115 |
| Diagnosis (svPPA) | -0.17 (0.41) | -0.98-0.64 | -0.41 | 0.684 |
| Voxel (SMC) | 2.58 (0.19) | 2.20-2.95 | 13.44 | < 0.001*** |
| **tCho (i.u.)** |  |  |  |  |
| Diagnosis (nfvPPA) | 0.11 (0.09) | -0.07-0.28 | 1.22 | 0.227 |
| Diagnosis (svPPA) | -0.12 (0.11) | -0.34-0.09 | -1.12 | 0.269 |
| Voxel (SMC) | -0.26 (0.05) | -0.35-(-0.16) | -5.38 | < 0.001*** |
| **tCr (i.u.)** |  |  |  |  |
| Diagnosis (nfvPPA) | 0.34 (0.18) | -0.02-0.69 | 1.84 | 0.071 |
| Diagnosis (svPPA) | 0.54 (0.23) | 0.10-0.99 | 2.40 | 0.019* |
| Voxel (SMC) | -0.40 (0.11) | -0.61-(-0.19) | -3.78 | < 0.001*** |
| **Glx (i.u.)** |  |  |  |  |
| Diagnosis (nfvPPA) | 0.91 (0.60) | -0.27-2.09 | 1.51 | 0.136 |
| Diagnosis (svPPA) | 0.01 (0.74) | -1.45-1.47 | 0.01 | 0.988 |
| Voxel (SMC) | -2.58 (0.33) | -3.23-(-1.93) | -7.79 | < 0.001*** |
| **GABA+ (i.u.)** |  |  |  |  |
| Diagnosis (nfvPPA) | -0.10 (0.28) | -0.65-0.45 | -0.37 | 0.716 |
| Diagnosis (svPPA) | 0.10 (0.36) | -0.60-0.80 | 0.28 | 0.782 |
| Voxel (SMC) | -0.11 (0.23) | -0.57-0.34 | -0.49 | 0.626 |

Comparing lvPPA and svPPA, the follow-up binary logistic regression model revealed a statistically significant association between diagnosis and tCr levels (odds ratio: 3.08; 95% confidence interval: 1.46-7.21; *p* = 0.005). Comparing lvPPA and nfvPPA, the association between diagnosis and tCr levels was not statistically significant (odds ratio: 1.72; 95% confidence interval: 1.00-3.12; *p* = 0.062). Including age and years since diagnosis in these models did not improve model fit (higher AIC and BIC), and these predictors were not statistically significant indicators of PPA subtype (*p* > 0.05 in all cases).

### 3.3 Regional Differences in Brain Metabolites and Voxel Composition

tNAA was lower in the IFG compared with the SMC (*p* < 0.001), while tCho, tCr, and Glx were higher in the IFG compared with the SMC (all *p* < 0.001; Figure 2; Table 2). GABA+ did not differ by brain region (*p* = 0.626; Figure 2; Table 2). The IFG compared with the SMC voxel had a higher gray matter (*p* < 0.001) and lower white matter fraction (*p* < 0.001), as well as a greater total tissue fraction (i.e., higher gray matter + white matter fraction; *p* = 0.012; Table B4).

### 3.4 Metabolite Relationships with PPA Symptom Severity

We included all short-TE PRESS metabolite levels from both voxels (i.e., 4 measures/voxel), age, sex, PPA subtype, and years since diagnosis in the full statistical model, and only those participants without missing short-TE PRESS or FTD-CDR data (*n* = 49; see Table B2). The final model retained left IFG tCr, left IFG Glx, and years since diagnosis as statistically significant predictors of FTD-CDR score (Table 3). That is, across the whole cohort, worse PPA symptoms (i.e., higher FTD-CDR scores) were associated with higher left IFG tCr levels (partial r = 0.39; *p* = 0.007; Figure 3), lower left IFG Glx levels (partial r = -0.36; *p* = 0.012; Figure 3), and longer disease duration (partial r = 0.34; *p* = 0.021). Rerunning this model with the square root of FTD-CDR scores as the outcome variable (to meet the normality assumption) returned the same three predictors and did not change the statistical significance of any results. Requiring that age and PPA subtype be included in the model did not change the statistical significance of the metabolite or disease duration predictors, and neither age (*p* = 0.529) nor PPA subtype (nfvPPA vs. lvPPA: *p* = 0.224; svPPA vs. lvPPA: *p* = 0.483) were significant predictors of FTD-CDR score.

**Table 3.** Metabolites Relationships with PPA Symptom Severity

Note.* Here we report the results of the final linear model testing for associations between brain metabolite levels and FTD-CDR (i.e., PPA symptom severity) scores. SE = standard error; CI = 95% confidence interval.
| Predictors | Estimates (SE) | CI | t | p | R<sup>2</sup> / R<sup>2</sup> Adjusted |
| --- | --- | --- | --- | --- | --- |
| (Intercept) | -14.18 (9.19) | -32.69-4.32 | -1.54 | 0.130 |  |
| IFG tCr (i.u.) | 1.96 (0.69) | 0.57-3.35 | 2.84 | **0.007\*\*** |  |
| IFG Glx (i.u.) | -0.55 (0.21) | -0.98-(-0.13) | -2.61 | **0.012\*** |  |
| Years since diagnosis | 0.48 (0.20) | 0.08-0.89 | 2.39 | **0.021\*** |  |
|  |  |  |  |  | 0.34 / 0.29 |

\* $p < 0.05$ , \*\* $p < 0.01$ .

**Figure 3.**
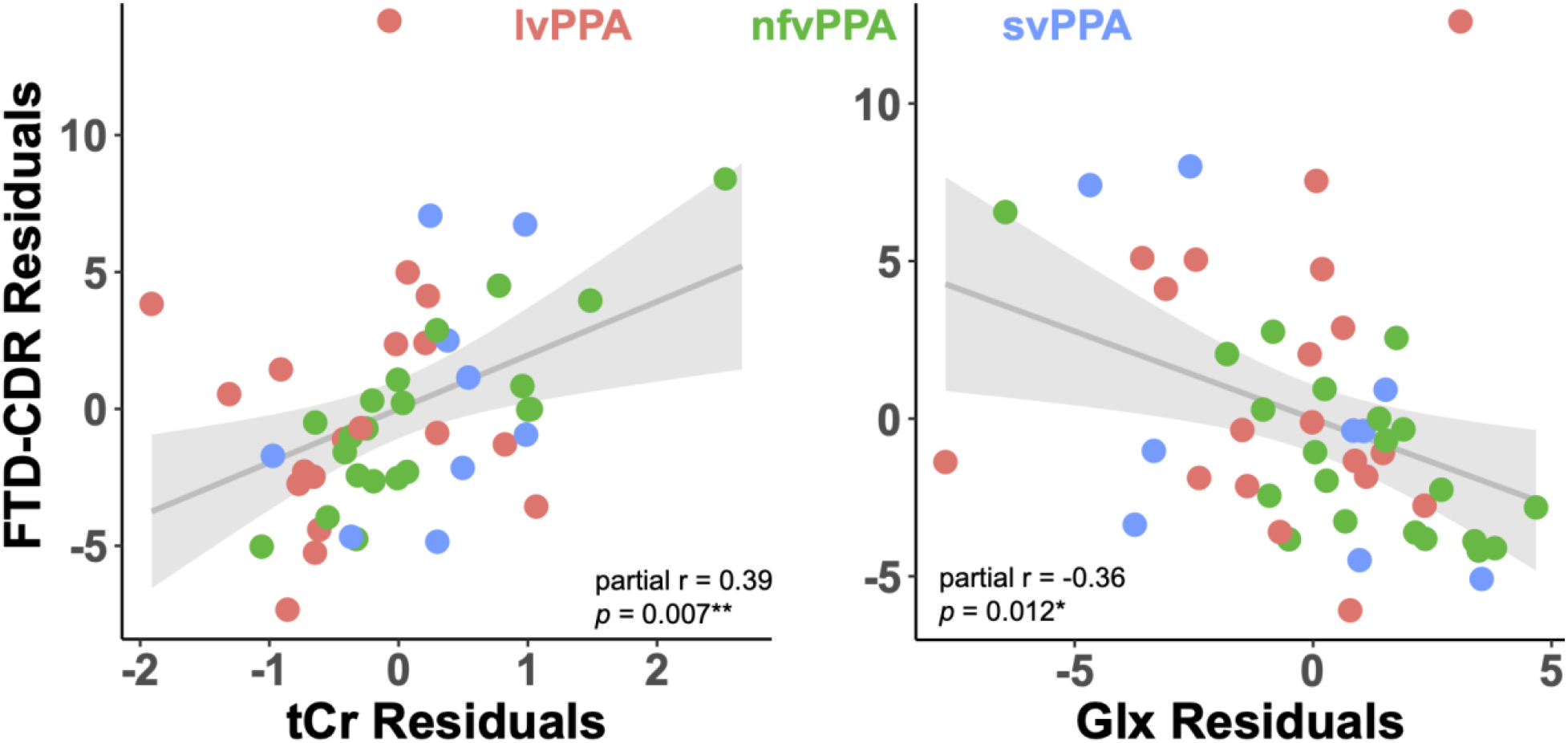
Relationship of IFG tCr and Glx with FTD-CDR Score. This figure depicts the partial correlations of tCr and Glx levels in the left IFG with FTD-CDR scores across all participants (lvPPA = pink, nfvPPA = green, svPPA = blue). These partial correlations account for the effects of the other predictors included in the final statistical model on the variable of interest (i.e., tCr values are corrected for Glx and years since diagnosis, and Glx values are corrected for tCr and years since diagnosis). Higher FTD-CDR scores indicate worse PPA symptom severity. These plots include *n* = 49 individuals, after exclusion of missing and unusable data (see Table B2).

## 4. Discussion

This is the first study to investigate brain metabolite differences between PPA subtypes using MRS. Our key findings included, firstly, that tissue-corrected tCr differed by PPA subtype across the left IFG and right SMC; lvPPA patients had the lowest tCr levels, followed by nfvPPA, and svPPA patients with the highest brain tCr levels. Moreover, tCr levels significantly differentiated between lvPPA and svPPA diagnosis. Secondly, higher tissue-corrected tCr and lower Glx levels in the left IFG (i.e., a brain region critical for language function) were associated with more severe symptoms as indicated by the FTD-CTR scale (which indexes language function as well as other cognitive and daily living domains (Knopman et al., 2008)). Together, these results suggest that brain tCr may play a role in PPA pathology or progression, particularly for the svPPA subtype, and could have utility for differentiating svPPA from other PPA subtypes. Both tCr and Glx might be explored as biomarkers for tracking disease severity and progression.

### 4.1 Higher tCr Levels in svPPA

Those with svPPA had the highest tCr levels across both the left IFG and right SMC. The creatine (Cr)/phosphocreatine (PCr) system is a short-lived energetic system that allows exchange of high-energy phosphate between Cr, adenosine triphosphate (ATP), and adenosine diphosphate (ADP) via creatine kinase, the primary enzyme in the Cr system. The Cr/PCr system simultaneously supplies a high-energy pool capable of rapidly regenerating ATP during high energetic demands, while also dispersing ATP across the cell to connect ATP consumption to the site of energy production (Bürklen et al., 2006). Cr is found in the highest concentrations in tissues with high ATP demand, including muscle and brain tissue (Rackayova et al., 2017; Rae, 2014). In the present work, we measured tCr (i.e., the sum of Cr and PCr); tCr is thus reflective of the Cr/PCr system more generally, including PCr anabolism, Cr degradation, and Cr extracellular transport.

Previous MRS studies have reported elevated tCr in normal aging (Chang et al., 1996; Charlton et al., 2007; Chiu et al., 2014; Cleeland et al., 2019; Haga et al., 2009; Pfefferbaum et al., 1999; Reyngoudt et al., 2012), MCI and AD (Huang et al., 2001), and other neurological conditions with cognitive symptoms (e.g., HIV-positive patients with neurological symptoms (Chang et al., 2003, 2002)). It remains unclear whether this is due to increased Cr and/or PCr synthesis, or decreased clearance of these metabolites from the brain (Chang et al., 1996). Elevated tCr in svPPA could also represent gliosis (Ratai et al., 2011; Reyngoudt et al., 2012; Saunders et al., 1999). tCr concentration in glial cells is about 2-4x that of neurons (Urenjak et al., 1993), and prior work has found an association between higher tCr in non-human primates and more severe gliosis (Ratai et al., 2011) and an association between PPA and accumulation of activated microglia in both gray (Kim et al., 2016) and white matter (Ohm et al., 2019). In addition, activated microglia have been reported to occur close to pathologic insults such as TDP43 inclusions in PPA patients with TDP43-positive pathology (Kim et al., 2016), i.e., the pathology most associated with svPPA (Gorno-Tempini et al., 2011).

It is also plausible that higher tCr in svPPA could be a compensatory response to reduced neuronal mitochondrial trafficking (i.e., movement of new mitochondria from the soma to distal axons (Cheng and Sheng, 2021)). Mitochondria are anchored in the cell by syntaphilin (SNPH) (Kang et al., 2008), which is elevated in TDP43-positive frontotemporal lobar degeneration (Andres-Benito et al., 2019), the pathology most associated with svPPA (Gorno-Tempini et al., 2011)). Elevated SNPH in svPPA could result in decreased mitochondrial motility, resulting in a greater need to disperse high-energy phosphate across the cell and thus elevated neuronal Cr. In support of this, neurons lacking appropriate mitochondrial trafficking machinery have reduced ability to form long dendrites, but this is rescued with Cr supplementation (Fukumitsu et al., 2015). Thus, while several mechanisms may contribute to the observed higher tCr levels in svPPA, the specific underlying cause requires further study.

### 4.2 Lower tCr Levels in lvPPA

It also remains unclear why tCr levels were the lowest for lvPPA patients. Though left IFG atrophy is reported to be more pronounced for lvPPA compared with svPPA (Preiß et al., 2019), we found tCr differences by PPA subtype across both left IFG and right SMC, suggesting more global tCr differences (not specific to one brain region). It is perhaps counterintuitive that tCr levels were lowest for lvPPA patients, given that lvPPA is most closely associated with AD pathology (Mesulam et al., 2008; Rabinovici et al., 2008) and tCr is elevated in AD compared with healthy controls (Huang et al., 2001). It is possible that the reported lvPPA tCr levels are in a similar range to those with AD. Though we are unaware of any studies which directly compare tCr levels in AD versus (lv)PPA patients, recent work found no differences in MRS-measured NAA and mI levels between AD and PPA patients (Mitolo et al., 2021), suggesting that certain brain metabolite levels might be similar between these two neurodegenerative conditions. However, without AD or control groups in the present work, we are unable to make such direct comparisons.

### 4.3 No PPA Subtype Differences for Other Neurometabolites

We identified differences in brain metabolite levels by PPA subtype only for tCr, but not for the other four investigated metabolites. The metabolite values reported here are corrected for bulk tissue concentrations (Gasparovic et al., 2006; Harris et al., 2015b); that is, we report metabolite values only within the brain tissue that remains in the voxels, accounting for age-related atrophy of gray and white matter. Therefore, our results are not likely explained entirely by subtype differences in atrophy location and severity. Moreover, as we did not identify PPA subtype differences in tNAA levels, it is less likely that tCr subtype differences were driven only by diffuse neuronal loss. Instead, we predict that these subtype differences in tCr are more related to differences in brain energy metabolism, though this hypothesis warrants further investigation in larger cohorts which include healthy aging controls.

### 4.4 tCr and Glx Associations with PPA Symptoms

We found that both higher tCr and lower Glx levels correlated with more severe PPA symptoms, as indicated by the FTD-CDR inventory. This suggests that higher brain tCr and lower brain Glx levels could be (directly or indirectly) related to maladaptive disease processes. These brain-behavior relationships occurred only for the left IFG but not the right SMC. The regional specificity of this association is plausible (and was hypothesized), as the FTD-CDR inventory includes both language and cognitive function sections; the left IFG is involved in language selection and production (e.g., both oral and written naming and spelling), as well as other cognitive functions (e.g., articulatory loop in working memory) (Liakakis et al., 2011), while the SMC roles are mostly related to motor execution and sensory processing. Higher tCr levels might indicate disruptions in underlying cellular bioenergetic processes, which could contribute to the development of more severe language and other cognitive symptoms in PPA patients; however, such claims warrant further replication in future studies with larger samples, as well as exploration of tCr relationships with additional cognitive metrics.

While higher tCr correlated with more severe PPA symptoms, higher brain tCr levels might not be entirely dysfunctional; the relationship between tCr and PPA disease processes could be more complex. Oral Cr supplementation has been shown to improve cognitive function in normal adults (Avgerinos et al., 2018) (though results differed by cognitive domain, effect sizes were small to moderate, and no large-scale Cr supplementation studies have been conducted in PPA patients). Moreover, multiple prior studies have reported *lower* tCr levels or no difference in tCr levels for normal aging cohorts (Cleeland et al., 2019). While these studies used varying methods (e.g., different MRI scanner field strengths and different brain regions), it remains unclear what other factors might influence brain tCr changes in aging and disease. Moreover, as we were unable to include an age-matched control group in the present study, it could be that the tCr levels we report for the PPA patients do not differ significantly from those of normal aging.

Prior work has identified relationships between lower Glx (or glutamate) and poorer cognitive and visuomotor performance in both normal aging (Levin et al., 2019; Zahr et al., 2013) and neurodegenerative disease (i.e., MCI (Oeltzschner et al., 2019; Zeydan et al., 2017) and frontotemporal lobar degeneration (Murley et al., 2022, 2020)). Reduced Glx could indicate loss of glutamatergic neurons or disturbances in glutamate and/or glutamine synthesis, which could each reasonably contribute to poorer functional performance. Moreover, as glutamate is a component involved in the synthesis of glutathione (i.e., one of the most abundant brain antioxidants), it could also be that increased brain oxidative stress in response to disease processes would result in greater glutathione production (Duffy et al., 2014; Hupfeld et al., 2021), and thus lower detectable brain glutamate levels. In our cohort, tCr was not correlated with Glx levels in either the IFG (Pearson r = -0.04; *p* = 0.782) or SMC (Pearson r = 0.13; *p* = 0.383), and thus each metabolite may distinctly contribute (directly or indirectly) to PPA disease processes. Further investigation is warranted to better understand the precise mechanisms by which tCr and Glx might each associate with PPA symptoms and progression.

### 4.5 Limitations

There are several limitations to this work. Our cross-sectional approach precluded us from assessing how these metabolite levels may alter prior to PPA onset and with disease progression. Though years since diagnosis did not differ between subtypes and was distinct from the tCr and Glx relationships with symptom severity, it still could be that disease progression rather than subtype more strongly influences brain metabolite levels. PPA is relatively rare and difficult to diagnose (Knopman et al., 2008); thus, we were unable to compare individuals at the same stage in their disease progression. Similarly, though age did not differ between subtypes and was distinct from the subtype differences in tCr levels, the reported tCr levels in PPA may not differ substantially from normal aging. We were unable to address this question due to the lack of a normal aging control group. There were no differences in the relationship between tCr and symptom severity based on PPA subtype, though all groups trended in the same direction; however, we were likely underpowered to investigate any group differences in this brain-behavior relationship.

In addition, there are several limitations to the MRS methods used. Our methods do not allow us to distinguish the contribution of PCr versus Cr to the tCr signal, or the contribution of glutamate versus glutamine to the Glx signal. The tissue corrections applied here are standard for aging cohorts with cortical atrophy (Gasparovic et al., 2006; Harris et al., 2015b); however, these corrections use literature values (Wansapura et al., 1999) for relaxation constants. Changes in water and metabolite relaxation rates with aging and disease could also impact metabolite quantification (Deelchand et al., 2020; Marjańska et al., 2017, 2013), though these were not quantified and thus could not be incorporated into our tissue corrections. The short-TE PRESS data were averaged on the scanner (instead of collecting individual spectra); therefore, frequency drift correction could not be performed on the short-TE PRESS data. It is recommended that future studies use post-processing drift corrections for all MRS data. PPA patients often have more difficulty than others remaining still in the scanner, which could have affected spectral quality (though we excluded any participants with NAA linewidth > 15 Hz); moreover, ability to complete an MRI scan precluded us from enrolling any extremely advanced PPA cases in this cohort.

## 5. Conclusions

We found that brain tCr differs between PPA subtypes. tCr is highest for svPPA patients, and brain tCr levels differentiate svPPA from lvPPA diagnosis. We also found a correlation between both higher left IFG tCr levels and lower left IFG Glx levels and more severe PPA symptoms. These results suggest that tCr may be useful in differentiating between PPA subtypes, and that both tCr and Glx could have utility as markers of PPA symptom severity. However, further work in larger cohorts which include a normal aging control group is needed to better understand the potential roles of tCr and Glx in PPA development and progression.

## Data Availability

The raw data supporting the conclusions of this manuscript will be made available by the authors without undue reservation.

## Ethics Approval and Consent to Participate

The Johns Hopkins University Institutional Review Board (#NA_00071337) approved all study procedures, and written informed consent was obtained from all participants.

## Consent for Publication

All authors consent to the publication of this study.

## Author Contributions

KH analyzed the MRS data, conducted all statistical analyses, created all figures and supplemental material, and wrote the first draft of the manuscript. HZ, GO, SH, AH, and RAE consulted on MRS data analyses and interpretation. HH contributed to manuscript writing and results interpretation. OH, JG, AH, and KT facilitated MRS and behavioral data collection. KT and RAE designed the project and led interpretation and discussion of the results. All authors participated in revision of the manuscript.

## Acknowledgements

The authors also wish to thank all of the participants who volunteered their time, without whom this project would not have been possible.

