## Appendix A, Appendix B, Appendix C for "Brain Total Creatine Differs Between Primary Progressive Aphasia (PPA) Subtypes and Correlates with Disease Severity"

### Appendix A: Summary MRSinMRS Report

Here we provide a summary following the minimum reporting standards in MRS generated in Osprey. For further details, see: Lin et al. 'Minimum Reporting Standards for *in vivo* Magnetic Resonance Spectroscopy (MRSinMRS): Experts' consensus recommendations. NMR in Biomedicine. 2021;e4484. doi.org/10.1002/nbm.4448

**Site:** The Johns Hopkins University School of Medicine / F. M. Kirby Research Center for Functional Brain Imaging, Kennedy Krieger Institute

| **1. Hardware** | |
|---|---|
| a. Field strength [T] | 3 T |
| b. Manufacturer | Philips |
| c. Model (software version if available) | R 5.1.7 |
| d. RF coils: nuclei (transmit/receive), number of channels, type, body part | 1H, 32 channel, head |
| e. Additional hardware | - |

| **2. Acquisition** | |
|---|---|
| **Short-TE PRESS** | |
| a. Pulse sequence | Philips PRESS |
| b. Volume of interest (VOI) locations | Left IFG, Right SMC |
| c. Nominal VOI size [$mm^3$] | 30 x 30 x 30 $mm^3$ |
| d. Repetition time (TR), echo time (TE) [ms] | TR 2000 ms, TE 32 ms |
| e. Total number of averages per spectrum | 48 total averages |
| f. Additional sequence parameters | F1: 2000 Hz, 2048 points |
| g. Water suppression method | VAPOR |
| h. Shimming method, reference peak, and threshold of acceptance of shim chosen | 2nd order pencil beam, water |
| i. Trigger or motion correction | No trigger or active motion correction |
| **MEGA-PRESS** | |
| a. Pulse sequence | MEGA-PRESS (Johns Hopkins University Patch) |
| b. Volume of interest (VOI) locations | Left IFG, Right SMC |
| c. Nominal VOI size [$mm^3$] | 30 x 30 x 30 $mm^3$ |
| d. Repetition time (TR), echo time (TE) [ms] | TR 2000 ms, TE 68 ms |
| e. Total number of averages per spectrum<br>i. Number of averaged spectra per subspectrum | 320 total averages with 160 averages per subspectrum |
| f. Additional sequence parameters | F1: 2000 Hz, 2048 points |
| i. editing pulse frequencies | $ppm_{ON}$ = 1.90, $ppm_{OFF}$ = 7.50 |
| g. Water suppression method | VAPOR |
| h. Shimming method, reference peak, and threshold of acceptance of shim chosen | 2nd order pencil beam, water |
| i. Trigger or motion correction | No trigger or active motion correction |

#### 3. Data analysis methods and outputs

| | |
|---|---|
| **Short-TE PRESS** | |
| a. Analysis software | Osprey 2.0.0 |
| b. Processing steps deviating from Osprey | None |
| c. Output measure | TissCorrWaterScaled (Gasparovic et al. 2006) |
| d. Quantification references and assumptions, fitting model assumptions | Basis set list: Asc,Asp,Cr,GABA,GPC,GSH,Gln,Glu,Ins,Lac, NAA,NAAG,PCh,PCr,PE,Scyllo,Tau,MM09,MM12, MM14,MM17,MM20,Lip09,Lip13,Lip20 Fitting method: Osprey baseline knot spacing 0.40 ppm |
| **MEGA-PRESS** | |
| a. Analysis software | Osprey 2.0.0 |
| b. Processing steps deviating from Osprey | Final alignment of the averaged sub-spectra by minimizing the choline (not water) peak; co-edited MMs at 3 ppm were modelled using the “1to1GABA” model |
| c. Output measure | AlphaCorrWaterScaled (Harris et al. 2015) |
| d. Quantification references and assumptions, fitting model assumptions | Basis set list: Asc,Asp,Cr,CrCH2,GABA,GPC,GSH,Gln,Glu,Ins, Lac,NAA,NAAG,PCh,PCr,PE,Scyllo,Tau,MM09, MM12,MM14,MM17,MM20,Lip09,Lip13,Lip20 Fitting method: Osprey baseline knot spacing 0.40 ppm |

#### 4. Data quality

| | |
|---|---|
| **Short-TE PRESS, Left IFG** | |
| a. SNR (NAA), linewidth (NAA) [Hz] | SNR: 135 ± 33, linewidth: 9.72 ± 1.78 Hz |
| b. Data exclusion criteria | NAA linewidth > 15 Hz |
| c. Quality measures of postprocessing model fitting (Mean Relative Amplitude Residual) | 3.36 % |
| d. Mean spectrum created with OspreyOverview | Figure 1B |
| **Short-TE PRESS, Right SMC** | |
| a. SNR (NAA), linewidth (NAA) [Hz] | SNR: 193 ± 32, linewidth: 8.20 ± 1.16 Hz |
| b. Data exclusion criteria | NAA linewidth > 15 Hz |
| c. Quality measures of postprocessing model fitting (Mean Relative Amplitude Residual) | 10.73 % |
| d. Mean spectrum created with OspreyOverview | Figure 1B |
| **GABA-Edited MEGA-PRESS, Left IFG** | |
| a. SNR (NAA), linewidth (NAA) [Hz, OFF spectra], avg. drift (Cr, mean (range)) [Hz, DIFF1 spectra] | SNR: 167 ± 51, linewidth: 7.31 ± 0.93 Hz, drift: 0.03 (-0.03-0.06) |
| b. Data exclusion criteria | NAA linewidth > 15 Hz |
| c. Quality measures of postprocessing model fitting (Mean Relative Amplitude Residual) | |
| OFF | 8.71 ± 9.33 |
| DIFF1 | 3.96 ± 1.58 |
| d. Mean spectrum created with OspreyOverview | Figure 1C |

| 4. Data quality | |
|---|---|
| **GABA-Edited MEGA-PRESS, Right SMC** | |
| a. SNR (NAA), linewidth (NAA) [Hz, OFF spectra], avg. drift (Cr, mean (range)) [Hz, DIFF1 spectra] | SNR: 211 ± 46, linewidth: 6.04 ± 0.71 Hz, drift: 0.05 (-0.05-0.09) |
| b. Data exclusion criteria | NAA linewidth > 15 Hz |
| c. Quality measures of postprocessing model fitting (Mean Relative Amplitude Residual) | |
| OFF | 17.80 ± 19.87 |
| DIFF1 | 2.97 ± 0.86 |
| d. Mean spectrum created with OspreyOverview | Figure 1C |

*Note.* NAA SNR is the maximum amplitude of the NAA peak divided by twice the standard deviation of the noise. The frequency drift over the course of each GABA-edited MEGA-PRESS experiment is determined based on the creatine signal at 3.02 ppm in each average. Mean drift is the average of the absolute value of change in the creatine signal for each participant across each experiment. Drift range is the average minimum and maximum value of change in the creatine signal for each participant across each experiment.

**Appendix B.** Supplemental Figures and Tables

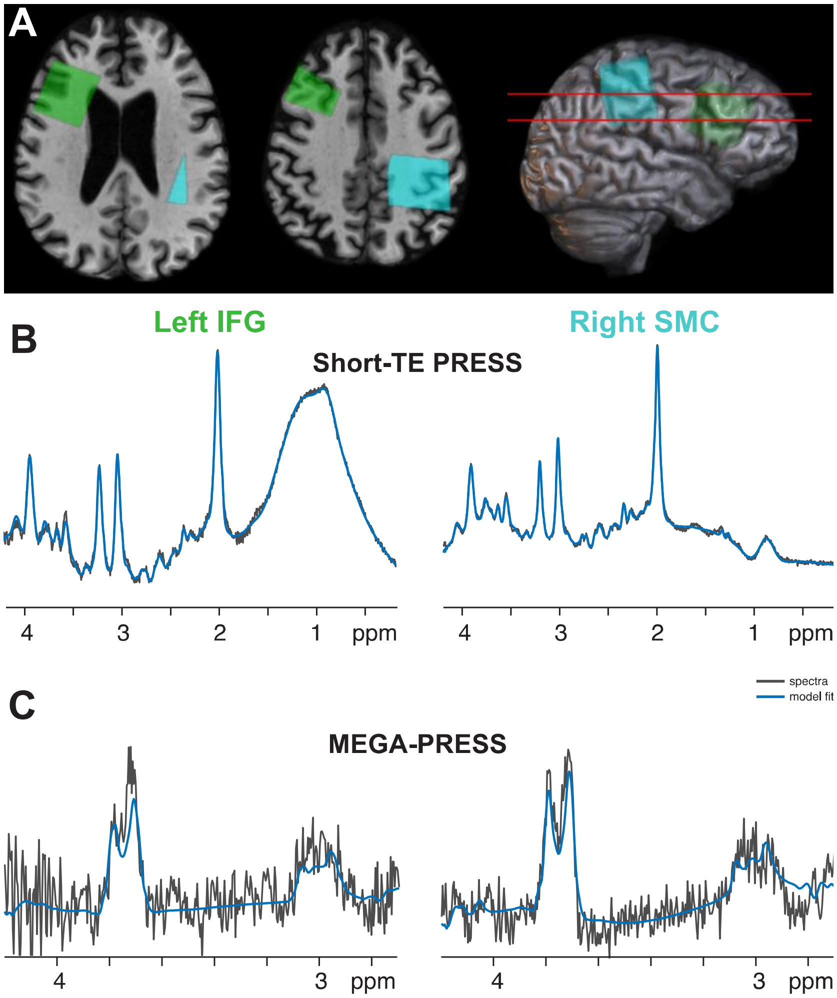

**Figure B1. Exemplar Voxel Placement and Spectra for One Participant. (A)** The placement of both voxels (IFG in green, SMC in blue) for a single exemplar participant is overlaid onto their skull-stripped structural image to protect patient privacy. The rendered structural image is shown semi-transparent, in order to depict voxel placement within the brain. **(B-C)** Here we depict the short-TE PRESS and GABA-edited MEGA-PRESS spectra (black) and model fits (blue) for the same exemplar participant for both voxels. The PRESS spectra are shown from 0.2-4.2 ppm. The MEGA-PRESS spectra are shown from 2.7-4.2 ppm. This representative participant was the individual with the median overall model fit (i.e., relative residual amplitude averaged across the four experiments).

**Table B1**. Data Quality by PPA Subtype

| | lvPPA | nfvPPA | svPPA | F | *p* | $\eta^2$ |
|---|---|---|---|---|---|---|
| **IFG short-TE PRESS** | | | | | | |
| SNR (NAA) [Hz] | 135.58 (34.33) | 135.96 (35.40) | 129.90 (25.11) | 0.15 | 0.861 | 0.01 |
| Linewidth (NAA) [Hz] [a] | 9.65 (1.91) | 9.82 (1.98) | 9.61 (9.32) | 0.07 | 0.929 | 0.003 |
| Model Fit (Mean Relative Amplitude Residual) | 3.14 (0.98) | 3.54 (1.33) | 3.38 (1.03) | 0.65 | 0.529 | 0.02 |
| **SMC short-TE PRESS** | | | | | | |
| SNR (NAA) [Hz] | 194.29 (19.92) | 189.57 (40.17) | 199.53 (29.98) | 0.42 | 0.661 | 0.01 |
| Linewidth (NAA) [Hz] [a] | 8.13 (1.19) | 8.33 (1.29) | 8.02 (0.83) | 0.34 | 0.711 | 0.01 |
| Model Fit (Mean Relative Amplitude Residual) [a] | 9.50 (7.22) | 13.58 (15.93) | 6.58 (2.35) | 1.72 | 0.189 | 0.06 |
| **IFG MEGA-PRESS** | | | | | | |
| SNR (NAA) [Hz] | 168.42 (48.66) | 169.86 (54.20) | 157.95 (49.97) | 0.18 | 0.836 | 0.01 |
| Linewidth (NAA) [Hz, OFF spectra] | 7.56 (0.92) | 7.27 (0.94) | 6.96 (0.94) | 1.28 | 0.289 | 0.05 |
| Drift (Cr, mean) [Hz, DIFF1 spectra] | 0.033 (0.015) | 0.032 (0.014) | 0.032 (0.012) | 0.024 | 0.976 | 0.001 |
| Model Fit (Mean Relative Amplitude Residual) [OFF spectra] | 7.13 (4.30) | 10.76 (12.80) | 6.43 (5.56) | 1.07 | 0.352 | 0.04 |
| Model Fit (Mean Relative Amplitude Residual) [DIFF1 spectra] | 4.16 (1.59) | 4.03 (1.62) | 3.41 (1.51) | 0.70 | 0.500 | 0.03 |
| **SMC MEGA-PRESS** | | | | | | |
| SNR (NAA) [Hz] | 205.65 (36.82) | 206.00 (50.78) | 233.71 (47.45) | 1.63 | 0.205 | 0.06 |
| Linewidth (NAA) [Hz, OFF spectra] [a] | 6.30 (0.80) | 5.96 (0.59) | 5.81 (0.73) | 2.10 | 0.133 | 0.07 |
| Drift (Cr, mean) [Hz, DIFF1 spectra] | 0.055 (0.019) | 0.053 (0.021) | 0.058 (0.022) | 0.21 | 0.810 | 0.01 |
| Model Fit (Mean Relative Amplitude Residual) [OFF spectra] [a] | 16.26 (16.85) | 21.79 (29.94) | 11.05 (3.03) | 1.23 | 0.302 | 0.04 |
| Model Fit (Mean Relative Amplitude Residual) [DIFF1 spectra] | 2.77 (0.72) | 3.19 (1.02) | 2.81 (0.57) | 1.62 | 0.208 | 0.06 |

*Table B1 Note*. For each metric, we report the mean (standard deviation) for each PPA subtype (after exclusion of the datasets noted in Table B2). We also report the results of one-way ANOVAs to characterize differences in each variable between the subtype groups.

[a] In cases where the variable did not meet the one-way ANOVA assumptions of homogeneity of variances between groups, independence of residuals, and/or normality of residuals (indicated by the subscript ‘a’), we reran each analysis using the non-parametric Kruskal-Wallis test. In all cases, the non-parametric test did not change the statistical significance of the result.

**Table B2**. Data Missing and Excluded from Statistical Analyses

| | **lvPPA** (total number of subjects) | **nfvPPA** (total number of subjects) | **svPPA** (total number of subjects) | **Reason for Exclusion** |
|---|---|---|---|---|
| Total *n* | 22 | 27 | 12 | -- |
| IFG short-TE PRESS | 20 | 24 | 12 | *n* = 2 missing<br>*n* = 3 NAA linewidth > 15 Hz |
| SMC short-TE PRESS | 22 | 27 | 12 | -- |
| IFG MEGA-PRESS | 17 | 23 | 9 | *n* = 1 missing<br>*n* = 6 NAA linewidth > 15 Hz<br>*n* = 5 incorrect sequence |
| SMC MEGA-PRESS | 19 | 26 | 11 | *n* = 5 incorrect sequence |
| FTD-CDR sum score | 22 | 23 | 9 | *n* = 7 missing |

**Table B3.** Differences in Metabolites by PPA Subtype and Brain Region

| Mean (SD) | | Predictors | Estimates (SE) | CI | t | *p* | Fit |
|---|---|---|---|---|---|---|---|
| **tNAA (i.u.)** | | | | | | | |
| lvPPA: 18.12 (1.85) | IFG: 16.51 (1.48) | **Fixed Effects** | | | | | |
| nfvPPA: 17.59 (2.03) | SMC: 19.05 (1.23) | *(Intercept)* | 16.75 (0.27) | 16.22-17.27 | 62.56 | **< 0.001***** | |
| svPPA: 17.87 (1.42) | | Diagnosis *(nfvPPA)* | -0.53 (0.33) | -1.19-0.12 | -1.60 | 0.115 | |
| | | Diagnosis *(svPPA)* | -0.17 (0.41) | -0.98-0.64 | -0.41 | 0.684 | |
| | | Voxel *(SMC)* | 2.58 (0.19) | 2.20-2.95 | 13.44 | **< 0.001***** | |
| | | **Random Effects** | | | | | |
| | | $\sigma^2$ | 1.02 | | | | |
| | | $\tau_{00\ \text{Participant}}$ | 0.76 | | | | |
| | | **Model Fit** | | | | | |
| | | Marginal $R^2$ | | | | | 0.49 |
| | | Conditional $R^2$ | | | | | 0.71 |
| **tCho (i.u.)** | | | | | | | |
| lvPPA: 2.94 (0.32) | IFG: 3.09 (0.33) | **Fixed Effects** | | | | | |
| nfvPPA: 3.04 (0.41) | SMC: 2.85 (0.38) | *(Intercept)* | 3.08 (0.07) | 2.94-3.22 | 44.03 | **< 0.001***** | |
| svPPA: 2.83 (0.33) | | Diagnosis *(nfvPPA)* | 0.11 (0.09) | -0.07-0.28 | 1.22 | 0.227 | |
| | | Diagnosis *(svPPA)* | -0.12 (0.11) | -0.34-0.09 | -1.12 | 0.269 | |
| | | Voxel *(SMC)* | -0.26 (0.05) | -0.35-(-0.16) | -5.38 | **< 0.001***** | |
| | | **Random Effects** | | | | | |
| | | $\sigma^2$ | 0.06 | | | | |
| | | $\tau_{00\ \text{Participant}}$ | 0.06 | | | | |
| | | **Model Fit** | | | | | |
| | | Marginal $R^2$ | | | | | 0.17 |
| | | Conditional $R^2$ | | | | | 0.57 |
| **tCr (i.u.)** | | | | | | | |
| lvPPA: 12.20 (0.70) | IFG: 12.65 (0.79) | **Fixed Effects** | | | | | |
| nfvPPA: 12.51 (0.84) | SMC: 12.27 (0.75) | *(Intercept)* | 12.41 (0.15) | 12.12-12.70 | 84.62 | **< 0.001***** | |
| svPPA: 12.75 (0.70) | | Diagnosis *(nfvPPA)* | 0.34 (0.18) | -0.02-0.69 | 1.84 | 0.071 | |
| | | Diagnosis *(svPPA)* | 0.54 (0.23) | 0.10-0.99 | 2.40 | **0.019*** | |
| | | Voxel *(SMC)* | -0.40 (0.11) | -0.61-(-0.19) | -3.78 | **< 0.001***** | |
| | | **Random Effects** | | | | | |
| | | $\sigma^2$ | 0.31 | | | | |
| | | $\tau_{00\ \text{Participant}}$ | 0.22 | | | | |
| | | **Model Fit** | | | | | |
| | | Marginal $R^2$ | | | | | 0.14 |
| | | Conditional $R^2$ | | | | | 0.49 |
| **Glx (i.u.)** | | | | | | | |
| lvPPA: 10.04 (2.68) | IFG: 11.81 (2.53) | **Fixed Effects** | | | | | |
| nfvPPA: 10.97 (2.79) | SMC: 9.22 (2.35) | *(Intercept)* | 11.40 (0.48) | 10.46-12.34 | 23.83 | **< 0.001***** | |
| svPPA: 10.12 (2.73) | | Diagnosis *(nfvPPA)* | 0.91 (0.60) | -0.27-2.09 | 1.51 | 0.136 | |
| | | Diagnosis *(svPPA)* | 0.01 (0.74) | -1.45-1.47 | 0.01 | 0.988 | |
| | | Voxel *(SMC)* | -2.58 (0.33) | -3.23-(-1.93) | -7.79 | **< 0.001***** | |
| | | **Random Effects** | | | | | |
| | | $\sigma^2$ | 3.03 | | | | |
| | | $\tau_{00\ \text{Participant}}$ | 2.58 | | | | |
| | | **Model Fit** | | | | | |
| | | Marginal $R^2$ | | | | | 0.38 |
| | | Conditional $R^2$ | | | | | NA |

| GABA+ (i.u.) | | | | | | | |
|---|---|---|---|---|---|---|---|
| lvPPA: 3.81 (1.32) | IFG: 3.83 (1.34) | **Fixed Effects** | | | | | |
| nfvPPA: 3.70 (1.24) | SMC: 3.72 (1.11) | *(Intercept)* | 3.87 (0.25) | 3.88-4.35 | 15.73 | **< 0.001*** ** | |
| svPPA: 3.89 (1.02) | | Diagnosis *(nfvPPA)* | -0.10 (0.28) | -0.65-0.45 | -0.37 | 0.716 | |
| | | Diagnosis *(svPPA)* | 0.10 (0.36) | -0.60-0.80 | 0.28 | 0.782 | |
| | | Voxel *(SMC)* | -0.11 (0.23) | -0.57-0.34 | -0.49 | 0.626 | |
| | | **Random Effects** | | | | | |
| | | $\sigma^2$ | 1.35 | | | | |
| | | $\tau_{00\ \text{Participant}}$ | 0.11 | | | | |
| | | **Model Fit** | | | | | |
| | | Marginal $R^2$ | | | | | 0.01 |
| | | Conditional $R^2$ | | | | | 0.08 |

*Table B3 Note*. *$p$ < 0.05, **$p$ < 0.01, ***$p$ < 0.001. SD = standard deviation; SE = standard error; CI = 95% confidence interval. On the left, we report the mean (standard deviation) for each metabolite, split by PPA subtype and by voxel. On the right, we report the results of a linear mixed effects model testing for PPA subtype and voxel differences in each metabolite. lvPPA and IFG served as the reference groups. Marginal $R^2$ values consider only the variance of the fixed effects; conditional $R^2$ values consider both the fixed and random effects.

**Table B4.** Differences in Tissue Fractions by PPA Subtype and Brain Region

| Mean (SD) | | Predictors | Estimates (SE) | CI | t | *p* | Fit |
|---|---|---|---|---|---|---|---|
| **Gray Matter Fraction** | | | | | | | |
| lvPPA: 0.31 (0.06) | IFG: 0.34 (0.05) | **Fixed Effects** | | | | | |
| nfvPPA: 0.29 (0.06) | SMC: 0.26 (0.04) | *(Intercept)* | 0.35 (0.01) | 0.33-0.37 | 42.98 | **< 0.001*** ** | |
| svPPA: 0.30 (0.06) | | Diagnosis *(nfvPPA)* | -0.02 (0.01) | -0.04-0.00 | -1.55 | 0.126 | |
| | | Diagnosis *(svPPA)* | -0.01 (0.01) | -0.04-0.01 | -0.85 | 0.397 | |
| | | Voxel *(SMC)* | -0.08 (0.01) | -0.10-(-0.07) | -13.95 | **< 0.001*** ** | |
| | | **Random Effects** | | | | | |
| | | $\sigma^2$ | 0.00 | | | | |
| | | $\tau_{00\ Participant}$ | 0.00 | | | | |
| | | **Model Fit** | | | | | |
| | | Marginal $R^2$ | | | | | 0.52 |
| | | Conditional $R^2$ | | | | | 0.71 |
| **White Matter Fraction** | | | | | | | |
| lvPPA: 0.54 (0.07) | IFG: 0.51 (0.07) | **Fixed Effects** | | | | | |
| nfvPPA: 0.54 (0.06) | SMC: 0.57 (0.05) | *(Intercept)* | 0.51 (0.01) | 0.48-0.53 | 43.25 | **< 0.001*** ** | |
| svPPA: 0.55 (0.07) | | Diagnosis *(nfvPPA)* | 0.001 (0.01) | -0.03-0.03 | 0.05 | 0.962 | |
| | | Diagnosis *(svPPA)* | 0.02 (0.02) | -0.02-0.05 | 0.98 | 0.329 | |
| | | Voxel *(SMC)* | 0.06 (0.01) | 0.05-0.08 | 8.11 | **< 0.001*** ** | |
| | | **Random Effects** | | | | | |
| | | $\sigma^2$ | 0.00 | | | | |
| | | $\tau_{00\ Participant}$ | 0.00 | | | | |
| | | **Model Fit** | | | | | |
| | | Marginal $R^2$ | | | | | 0.24 |
| | | Conditional $R^2$ | | | | | 0.61 |
| **Total Tissue (Gray Matter Fraction + White Matter Fraction)** | | | | | | | |
| lvPPA: 0.84 (0.05) | IFG: 0.85 (0.06) | **Fixed Effects** | | | | | |
| nfvPPA: 0.83 (0.06) | SMC: 0.83 (0.05) | *(Intercept)* | 0.86 (0.01) | 0.83-0.88 | 80.96 | **< 0.001*** ** | |
| svPPA: 0.85 (0.05) | | Diagnosis *(nfvPPA)* | -0.01 (0.01) | -0.04-0.01 | -1.12 | 0.268 | |
| | | Diagnosis *(svPPA)* | 0.01 (0.02) | -0.02-0.04 | 0.46 | 0.645 | |
| | | Voxel *(SMC)* | -0.02 (0.01) | -0.04-(-0.01) | -2.61 | **0.012*** | |
| | | **Random Effects** | | | | | |
| | | $\sigma^2$ | 0.00 | | | | |
| | | $\tau_{00\ Participant}$ | 0.00 | | | | |
| | | **Model Fit** | | | | | |
| | | Marginal $R^2$ | | | | | 0.06 |
| | | Conditional $R^2$ | | | | | 0.42 |

*Table B4 Note*. *$p < 0.05$, **$p < 0.01$, ***$p < 0.001$. SD = standard deviation; SE = standard error; CI = 95% confidence interval. On the left, we report the mean (standard deviation) for each tissue fraction, split by PPA subtype and by voxel. On the right, we report the results of a linear mixed effects model testing for PPA subtype and voxel differences in each tissue fraction. lvPPA and IFG served as the reference groups. Marginal $R^2$ values consider only the variance of the fixed effects; conditional $R^2$ values consider both the fixed and random effects.

**Appendix C.** Quantification and Tissue Correction Details

Here we provide further details regarding the metabolite quantification and tissue correction conduced within the Osprey package. For further details regarding Osprey, please refer to (Oeltzschner et al., 2020). For all Osprey source code and documentation, please visit: https://github.com/schorschinho/osprey/

**Water Scaling in Osprey:**

As described in (Oeltzschner et al., 2020), Osprey reports water-scaled metabolite estimates when an unsuppressed water signal is provided. Osprey reports water-scaled metabolite estimates according to Equation 1. This is analogous to the LCModel water scaling procedure, which does not account for tissue composition and assumes pure white matter by default.

**Equation 1.** Water Scaling in Osprey

$$[met]H_2O = \frac{S_{met}}{S_{H_2O}} \text{ x } [H_2O] \text{ x } conc_{H_2O} \text{ x } \frac{1 - e^{-\frac{TR_w}{T_{1w}}}}{1 - e^{-\frac{TR_{met}}{T_{1met}}}} \text{ x } \frac{e^{-\frac{TE_w}{T_{2w}}}}{e^{-\frac{TE_{met}}{T_{2met}}}}$$

In Equation 1:

- $S_{met}$ and $S_{H_2O}$ are metabolite and water signal amplitudes.
- $[H_2O]$ is the molal concentration of pure MR-visible water, i.e., 55.5 mol/kg of MR-visible water (Gasparovic et al., 2006; Knight-Scott et al., 2003).
- $conc_{H_2O}$ is the relative water density of white matter, i.e., 0.65.
- $TR_w$, $TR_{met}$, $TE_w$, $TE_{met}$ are the repetition times (TR) and echo times (TE) of the water-unsuppressed and water-suppressed acquisitions.
- $T_{1w}$ and $T_{2w}$ are the averaged relaxation times for tissue water. For brain data at 3 Tesla, $T_{1w}$ = 1100 ms and $T_{2w}$ = 95 ms (Wansapura et al., 1999).
- $T_{1met}$ and $T_{2met}$ are the averaged relaxation times of all metabolites and generated from a lookup table.

**Short-TE PRESS Data: Gasparovic Tissue Correction**

The short-TE PRESS metabolite values included in all statistical models were tissue-corrected. As described in (Oeltzschner et al., 2020), Osprey calculates fully tissue- and relaxation-corrected molal concentration estimates according to Equation 2, the Gasparovic method (Gasparovic et al., 2006).

**Equation 2.** Gasparovic Tissue Correction

$$[met]TissCorr = \frac{S_{met}}{S_{H_2O}(1 - f_{CSF})} \text{ x } [H_2O] \text{ x } \frac{f_{GM} \text{ x } R_{H_2O\_GM} + f_{WM} \text{ x } R_{H_2O\_WM} + f_{CSF} \text{ x } R_{H_2O\_CSF}}{R_{met}}$$

In Equation 2:

- $S_{met}$ and $S_{H_2O}$ are metabolite and water signal amplitudes.
- $f_{GM}$, $f_{WM}$, and $f_{CSF}$ are the molal water fractions for GM, WM and CSF, which are derived from the volume fractions according to Equation 3.
- $R_{H_2O\_GM}$ , $R_{H_2O\_WM}$ and $R_{H_2O\_CSF}$ tissue-specific relaxation corrections are calculated according to Equation 4.

**Equation 3.** Molal Water Fractions for GM, WM, and CSF.

$$f_k = \frac{f_{vol,k} \text{ x } conc_{H_2O,k}}{f_{vol,GM} \text{ x } conc_{H_2O,GM} + f_{vol,WM} \text{ x } conc_{H_2O,WM} + f_{vol,CSF} \text{ x } conc_{H_2O,CSF}}$$

In Equation 3, the relative water densities are:

- $conc_{H_2O,GM}$ = 0.78$[H_2O]$
- $conc_{H_2O,WM}$ = 0.65$[H_2O]$
- $conc_{H_2O,CSF}$ = 0.97$[H_2O]$

**Equation 4.** Tissue-Specific Relaxation Corrections

$$R_{H_2O_k} = \left(1 - e^{-\frac{TR_w}{T_{1w\_k}}}\right) \text{ x } e^{-\frac{TE_w}{T_{2w\_k}}}$$

$$R_{met} = \left(1 - e^{-\frac{TR_{met}}{T_{1met}}}\right) \text{ x } e^{-\frac{TE_{met}}{T_{2met}}}$$

In Equation 4, the relaxation times for water and the metabolites at 3 Tesla were adapted from several widely-cited references (Edden et al., 2012; Mlynárik et al., 2001; Puts et al., 2013; Wansapura et al., 1999; Wyss et al., 2018). These values are listed in Tables C1-C2 and are available in the OspreyQuantify.m function within the Osprey package (https://github.com/schorschinho/osprey/).

**Table C1**. Water Relaxation Times Used in Gasparovic Tissue Correction – Equation 4

| | **Time (s)** |
|---|---|
| $T1_{w\_GM}$ | 1.331 |
| $T2_{w\_GM}$ | 0.110 |
| $T1_{w\_WM}$ | 0.832 |
| $T2_{w\_WM}$ | 0.0792 |
| $T1_{w\_CSF}$ | 3.817 |
| $T2_{w\_CSF}$ | 0.503 |

**Table C2**. Metabolite Relaxation Times Used in Gasparovic Tissue Correction – Equation 4

| | **Time (ms)** |
|---|---|
| **tNAA** | |
| $T1_{tNAA\_GM}$ | 1405 |
| $T2_{tNAA\_GM}$ | 187.75 |
| $T1_{tNAA\_WM}$ | 1270 |
| $T2_{tNAA\_WM}$ | 264 |
| **tCho** | |
| $T1_{tCho\_GM}$ | 1190 |
| $T2_{tCho\_GM}$ | 247.75 |
| $T1_{tCho\_WM}$ | 1080 |
| $T2_{tCho\_WM}$ | 215.5 |
| **tCr** | |
| $T1_{tCr\_GM}$ | 1460 |
| $T2_{tCr\_tGM}$ | 146 |
| $T1_{tCr\_WM}$ | 1240 |
| $T2_{tCr\_WM}$ | 166 |
| **Glx** | |
| $T1_{Glx\_GM}$ | 1305 |
| $T2_{Glx\_tGM}$ | 119.5 |
| $T1_{Glx\_WM}$ | 1180 |
| $T2_{Glx\_WM}$ | 146 |

#### GABA-Edited MEGA-PRESS Data: Alpha Correction

The GABA-edited MEGA-PRESS GABA+ values included in all statistical models were tissue-corrected. As described in (Harris et al., 2015), for GABA-edited spectra, Osprey calculates an additional 'alpha' correction metric which normalizes for the fact that GABA levels are higher in gray matter than in white matter (Jensen et al., 2005). For further details, see Equation 5 in (Harris et al., 2015) and the OspreyQuantify.m function within the Osprey package (https://github.com/schorschinho/osprey/).

#### Appendix C References

Edden, RAE, Intrapiromkul, J, Zhu, H, Cheng, Y, Barker, PB, 2012. Measuring T2 in vivo with J-difference editing: Application to GABA at 3 tesla. J. Magn. Reson. Imaging 35, 229–234. https://doi.org/10.1002/jmri.22865

Gasparovic, C, Song, T, Devier, D, Bockholt, HJ, Caprihan, A, Mullins, PG, Posse, S, Jung, RE, Morrison, LA, 2006. Use of tissue water as a concentration reference for proton spectroscopic imaging. Magn. Reson. Med. Off. J. Int. Soc. Magn. Reson. Med. 55, 1219–1226.

Harris, AD, Puts, NA, Edden, RA, 2015. Tissue correction for GABA-edited MRS: Considerations of voxel composition, tissue segmentation, and tissue relaxations. J. Magn. Reson. Imaging 42, 1431–1440.

Jensen, JE, deB. Frederick, B, Renshaw, PF, 2005. Grey and white matter GABA level differences in the human brain using two-dimensional,J-resolved spectroscopic imaging. NMR Biomed. 18, 570–576. https://doi.org/10.1002/nbm.994

Knight-Scott, J, Haley, AP, Rossmiller, SR, Farace, E, Mai, VM, Christopher, JM, Manning, CA, Simnad, VI, Siragy, HM, 2003. Molality as a unit of measure for expressing 1H MRS brain metabolite concentrations in vivo. Magn. Reson. Imaging 21, 787–797. https://doi.org/10.1016/S0730-725X(03)00179-6

Mlynárik, V, Gruber, S, Moser, E, 2001. Proton $T_1$ and $T_2$ relaxation times of human brain metabolites at 3 Tesla: METABOLITE $T_1$ AND $T_2$ IN HUMAN BRAIN AT 3 T. NMR Biomed. 14, 325–331. https://doi.org/10.1002/nbm.713

Oeltzschner, G, Zöllner, HJ, Hui, SC, Mikkelsen, M, Saleh, MG, Tapper, S, Edden, RA, 2020. Osprey: Open-source processing, reconstruction & estimation of magnetic resonance spectroscopy data. J. Neurosci. Methods 343, 108827.

Puts, NAJ, Barker, PB, Edden, RAE, 2013. Measuring the longitudinal relaxation time of GABA in vivo at 3 tesla. J. Magn. Reson. Imaging 37, 999–1003. https://doi.org/10.1002/jmri.23817

Wansapura, JP, Holland, SK, Dunn, RS, Ball Jr, WS, 1999. NMR relaxation times in the human brain at 3.0 tesla. J. Magn. Reson. Imaging Off. J. Int. Soc. Magn. Reson. Med. 9, 531–538.

Wyss, PO, Bianchini, C, Scheidegger, M, Giapitzakis, IA, Hock, A, Fuchs, A, Henning, A, 2018. In vivo estimation of transverse relaxation time constant ($T_2$) of 17 human brain metabolites at 3T: $T_2$ of 17 Human Brain Metabolites at 3T. Magn. Reson. Med. 80, 452–461. https://doi.org/10.1002/mrm.27067
